# The benefits of investments to combat HIV, tuberculosis and malaria for primary health care, 2000–2023: an economic modeling analysis

**DOI:** 10.1101/2025.05.02.25326907

**Authors:** Jiaying Stephanie Su, John Stover, Carel Pretorius, Peter Winskill, Sedona Sweeney, Timothy B. Hallett, Nicolas A. Menzies

**Affiliations:** Department of Global Health and Population, Harvard T.H. Chan School of Public Health, Boston, MA, USA; Center for Health Decision Science, Harvard T.H. Chan School of Public Health, Boston, MA, USA; Avenir Health, Glastonbury, Connecticut, USA; MRC Centre for Global Infectious Disease Analysis, School of Public Health, Imperial College London, London, UK; Department of Global Health and Development, Faculty of Public Health Policy, London School of Hygiene and Tropical Medicine, London, UK

## Abstract

**Background:** Global investments to combat HIV, tuberculosis, and malaria (HTM) have delivered substantial health gains, and may have reduced the burden placed by these diseases on the routine health system. We estimated the reduction in primary health care (PHC) utilization resulting from the scale-up of HTM services over 2000–2023 in 108 low- and middle-income countries.

**Methods:** For each disease, we applied established mathematical models to quantify PHC utilization (outpatient visits and inpatient bed-days provided outside of HTM programs) by individuals with symptomatic HIV, tuberculosis, or malaria unable to access HTM-specific services. For each country, we estimated averted PHC utilization by comparing a scenario describing the actual scale-up of HTM services to a counterfactual scenario holding HTM service coverage constant at year 2000 levels. We applied published unit costs to estimate the averted costs resulting from reduced PHC utilization.

**Findings:** Over 2000–2023, scale-up of HTM services averted an estimated 6·9 (95% interval: 4·4–10·4) billion outpatient PHC visits and 3·9 (2·5–6·1) billion inpatient bed-days, representing US$135 (70–248) billion in averted costs. These reductions were greatest in Sub-Saharan Africa and East Asia and Pacific regions. Across study countries, these reductions represented a median of 4·4% of hospital bed capacity and 1·6% of government health spending in 2023. These percentages were 22·9% and 5·1% respectively for low-income countries.

**Interpretation:** Over recent decades, sustained investments in HTM services in high-burden settings have averted substantial PHC utilization and associated costs. These benefits should be considered when assessing investment impact.

**Funding:** The Global Fund.

## Introduction

Since 2000, global efforts to combat HIV, tuberculosis (TB), and malaria have reduced the incidence of HIV by 54%, TB by 26%, and malaria by 24%.^1–3^ These achievements would not have been possible without sustained investments to scale up services to prevent and treat these diseases. These investments have been made by domestic governments and international partners, with funding provided through bilateral mechanisms such as the U.S. President’s Emergency Plan for AIDS Relief and multilateral mechanisms such as the Global Fund to Fight AIDS, Tuberculosis and Malaria. In 2023, approximately US$30 billion was spent by both domestic and international partners to advance HIV, TB, and malaria elimination in low- and middle-income countries (LMICs),^1–3^ reflecting global commitment to ending these diseases as public health threats.

Analyses of investments to address HIV, TB, and malaria (HTM) have typically compared the health benefits of these interventions to the resources required to scale and maintain HTM-specific services.^4–6^ However, less attention has been directed toward the implications for primary health care (PHC), except when these services are involved in providing HTM services. Although normative guidelines for economic evaluation stress the value of considering all consequences,^7^ evidence regarding the impact of HTM investments on health systems remains inconclusive. Concerns have been raised that global health initiatives may distort incentives within the health system and undermine services for non-targeted conditions, while reviews of empirical studies have documented both positive and negative effects.^8,9^ These studies highlight the need to quantify the implications of disease-specific investments on health systems, comparing possible outcomes under different levels of disease-specific investment. One way in which HTM investments could impact health systems is through changes in demand for PHC services. These changes in demand can arise through multiple mechanisms. First, disease-specific care funded through HTM programs will reduce the number of individuals experiencing untreated symptomatic disease, potentially crowding out care that would otherwise be sought from PHC providers. Second, improved HTM care prevents progression to advanced illness, reducing the need for inpatient care. Third, better HTM prevention and treatment can interrupt disease transmission, preventing new infections so that fewer symptomatic individuals require care at all. Each of these mechanisms could reduce PHC demand and free up health care resources for other uses, which would be particularly valuable in countries with limited health care resources.

In the context of evolving global priorities and growing fiscal constraints, it is important to understand not only the direct outcomes of disease-specific investments but also their broader implications for health system capacity and expenditure. In this study, we estimated the impact of investments to scale-up HIV, TB, and malaria services over 2000–2023 on primary health care utilization and costs outside of HTM-specific programs. We conducted this analysis for 108 LMICs with a high burden of HIV, TB, or malaria.

## Methods

### Population and analytic scenarios

We considered individuals with or at risk of HIV, TB, or malaria in LMICs with a significant burden of one of these conditions, and for which epidemiological outcomes were available from validated disease simulation models for each disease.^10–12^ We included 108 countries for HIV, 51 countries for TB, and 55 countries for malaria in our analyses, representing 94% of global HIV prevalence, 92% of global TB incidence, and 90% of global malaria incidence in 2023.

For each disease, we compared two scenarios covering years 2000–2023: an *actual scenario* reflecting the observed scale-up of HTM services in each country since year 2000, and a counterfactual *constant coverage scenario* holding HTM service coverage fixed at year 2000 levels. Changes in PHC utilization and costs were estimated from the difference between these scenarios in the number of individuals with symptomatic HIV, tuberculosis, or malaria not receiving effective disease-specific care through HTM programs. In sensitivity analyses, we assessed an additional counterfactual (*no coverage scenario)* in which HTM service coverage was reduced to zero from 2000 onward throughout the study period. Scenarios were assumed to be identical prior to 2000.

### Estimation of disease epidemiology

We obtained epidemiological outcomes for each disease and scenario from models employed for the Global Fund 8^th^ replenishment investment case.^13^ These models included the *Goals Model* for HIV, the *Global Portfolio TB Model* for TB, and the *malariasimulation* model for malaria.^10–12^ For the *actual scenario*, models incorporated the historical scale-up of HTM services (defined as the fraction of eligible individuals receiving each service) as inputs, and were calibrated to epidemiological estimates published by WHO and UNAIDS.^2,3,14^ Counterfactual scenarios represented hypothetical changes in HTM service coverage over the study period, with all other modeled mechanisms (e.g., demographic processes, intervention quality) following the assumptions of the *actual scenario*. For TB, we used the modeled results from the *Global Portfolio TB Model* (29 countries) to train a regression-based emulator and simulate epidemiological outcomes for an additional 22 high TB burden countries (details in Supplement).^2^ This increased the fraction of global TB incidence represented in the analysis from 81% to 92%.

### Estimation of averted PHC utilization

We used the modeled epidemiological estimates to calculate the number of individuals with symptomatic conditions due to HIV, TB, or malaria unable to access effective HTM-specific care. For HIV, this was operationalized as the number of person-years lived by persons with HIV not receiving antiretroviral therapy (ART). For TB and malaria, this was operationalized as the number of persons developing each disease who do not receive effective disease-specific care. For each disease, we computed the differences in these quantities between the actual and counterfactual scenarios over the study period. These differences were used to quantify averted PHC utilization associated with the scale-up of HTM services, which is not normally estimated by other analyses. To calculate the averted utilization, we applied estimates of the additional outpatient care (number of outpatient clinic visits) and inpatient care (number of hospital bed-days) received by affected individuals, as compared to individuals not experiencing these conditions. We did not include the utilization associated with the provision of HTM-specific diagnosis and treatment, as other analyses of HTM-specific services cover these quantities. We conducted the analysis separately for each country and disease. Values and sources for study inputs are provided in Supplement.

*HIV*: We computed the annual difference in the number of person-years spent by persons with HIV not on ART between scenarios, stratified into health states defined by age group (0-4 years, 5-14 years, and ≥15 years) and CD4 cell count category (<50, 50-99, 100-199, 200-249, 250-349, 350-499, and ≥500 cells/mm³). To estimate averted PHC outpatient visits, we multiplied this difference by the additional outpatient utilization per person-year for each health state, as compared to HIV-negative individuals (Figure S-1a, Table S-1a). The additional outpatient visits per person with HIV per year were estimated using rate ratios of outpatient visits for persons with HIV not on ART relative to HIV-negative individuals, and country-specific estimates of outpatient utilization in the general population. To estimate averted inpatient utilization, we multiplied the difference in the number of person-years of individuals with HIV not on ART by the additional annual hospitalizations associated with these individuals and the average duration of hospital stay.

*TB*: We calculated the annual difference in TB incidence between scenarios, stratified by whether or not these individuals received effective TB treatment. For those receiving treatment, we allowed for additional PHC utilization during the period before TB diagnosis and treatment, multiplying the difference in the number of new TB cases receiving effective treatment by the additional outpatient visits per TB case (Figure S-1b, Table S-1b). For untreated cases, we applied a number of additional outpatient visits over the whole disease episode (i.e., until death or self-cure) to the difference in the number of new TB cases not receiving effective treatment. We summed values for treated and untreated TB to derive the total averted outpatient utilization. The averted inpatient utilization was computed by multiplying the difference in the number of new TB cases not receiving effective treatment by the proportion hospitalized (conservatively assumed to be restricted to those dying with TB without TB treatment) and the average duration of hospitalization.

*Malaria*: We calculated the annual number of averted outpatient visits by multiplying the difference in the number of individuals with symptomatic malaria not receiving effective care between scenarios by the PHC utilization rates for individuals with symptomatic malaria, adjusted for country-specific differences in access to care (Figure S-1c, Table S-1c). For averted inpatient utilization, we applied the probabilities of hospitalizations for uncomplicated and severe malaria to the differences in the number of individuals with respective malaria conditions. We summed these results, subtracted out those who were hospitalized with malaria and received effective treatment, then multiplied the resulting estimate by the average duration of hospitalization to obtain the estimates of averted inpatient bed-days.

### Estimation of averted PHC costs

We estimated the averted costs associated with reduced PHC utilization by applying country-specific unit costs to the estimates of averted outpatient visits and inpatient bed-days. Unit costs were retrieved from WHO-CHOICE estimates and adjusted for inflation to reflect nominal U.S. dollars by year.^15–17^

### Outcomes

Primary outcomes are the averted PHC outpatient visits, PHC inpatient bed-days, and PHC costs resulting from the scale-up of HTM services over the study period. Results are reported by disease, country, year, World Bank region, and income classification. To contextualize these findings, we expressed averted inpatient utilization as a percentage of national hospital bed capacity, and averted PHC costs as a percentage of domestic government health expenditures.^18,19^

### Sensitivity analyses

We used probabilistic sensitivity analysis to quantify uncertainty in analytic outcomes.^20^ We specified probability distributions for each uncertain parameter (Table S-1), and drew 1000 parameter samples using a Latin hypercube design. We used these samples to generate 1000 estimates for each study outcome and calculated equal-tailed 95% uncertainty intervals from the distribution of results. We calculated partial rank correlation coefficients (PRCCs) to assess the relative importance of individual parameters in determining total averted costs.^21^ We also conducted sensitivity analyses around the cost functions used to estimate averted costs, testing non-linear cost functions to account for economies or diseconomies of scale for PHC services under the counterfactual scenarios (details in Supplement). All analyses were conducted in R (v4.4.1).

## Results

### Freed-up primary health care resources

Over 2000–2023, we estimated substantial reductions in PHC utilization due to the scale-up of HIV, TB, and malaria services across the 108 LMICs included in the analysis. In total, 6·9 (95% uncertainty interval: 4·4–10·4) billion outpatient PHC visits and 3·9 (2·5–6·1) billion inpatient bed-days were averted (Table 1). This reduction in utilization was equivalent to US$135 (70–248) billion in averted costs. In the Sub-Saharan Africa (SSA) region, where reductions in utilization were greatest for HIV and malaria, the scale-up of HIV services was estimated to have averted 0·9 (0·6–1·3) billion outpatient visits and 1·1 (0·4–2·2) billion inpatient bed-days over the study period, translating into US$43 (13–113) billion in averted costs (Table S-2). For malaria, the scale-up of services in SSA was estimated to have averted 2·3 (1·3–3·6) billion outpatient visits and 0·3 (0·2–0·6) billion inpatient bed-days, corresponding to US$4·9 (2·8–7·9) billion in averted costs. Scale-up of TB services contributed a notable portion of utilization reductions in the East Asia and Pacific region, averting 1·7 (0·6–3·6) billion outpatient visits and 1·0 (0·5–1·8) billion inpatient bed-days, representing US$53 (19–119) billion in averted costs between 2000 and 2023.

**Table 1.** Averted primary health care utilization and costs due to scale-up HIV, TB, and malaria services, for actual scenario compared to constant coverage scenario, 2000-23. Values in parentheses represent 95% uncertainty intervals. Cost values represent nominal U.S. dollars. Actual scenario represents the observed scale-up of HIV, TB, and malaria services over the study period. Constant coverage scenario represents a counterfactual with coverage of HIV, TB, and malaria services held constant at the year 2000 levels for each country over the study period.

| | Averted utilization, millions | | Averted costs (US\$), millions | | |
| --- | --- | --- | --- | --- | --- |
|  | Outpatient visits | Inpatient bed-days | Outpatient | Inpatient | Total |
| Total | 6926<br>(4362, 10447) | 3896<br>(2465, 6097) | 20547<br>(9150, 44503) | 114360<br>(52155, 226693) | 134906<br>(70124, 247531) |
| Disease |  |  |  |  |  |
| HIV | 1188<br>(757, 1734) | 1352<br>(516, 2710) | 4041<br>(1927, 7677) | 50121<br>(14679, 124034) | 54161<br>(17960, 129606) |
| TB | 3303<br>(1127, 6816) | 2150<br>(1164, 3579) | 13447<br>(3357, 36657) | 61431<br>(24074, 131111) | 74878<br>(35791, 148883) |
| Malaria | 2435<br>(1371, 3872) | 395<br>(184, 767) | 3058<br>(1455, 5423) | 2808<br>(1186, 6041) | 5866<br>(3344, 9539) |
| World Bank region |  |  |  |  |  |
| East Asia and Pacific | 1837<br>(705, 3751) | 1129<br>(621, 1856) | 10392<br>(1642, 33250) | 48983<br>(16114, 115361) | 59375<br>(22689, 127817) |
| Europe and Central Asia | 207<br>(87, 398) | 95<br>(57, 149) | 910<br>(305, 2156) | 3417<br>(1708, 6139) | 4327<br>(2315, 7431) |
| Latin America and Caribbean | 245<br>(134, 421) | 156<br>(89, 256) | 815<br>(305, 1774) | 4359<br>(1759, 8689) | 5174<br>(2444, 9764) |
| Middle East and North Africa | 5.2<br>(3.5, 7.4) | 3.8<br>(1.6, 7.3) | 28<br>(12, 55) | 169<br>(59, 385) | 197<br>(80, 421) |
| South Asia | 791<br>(364, 1419) | 594<br>(356, 936) | 1249<br>(259, 3286) | 5886<br>(2076, 13093) | 7136<br>(2781, 14502) |
| Sub-Saharan Africa | 3841<br>(2592, 5355) | 1918<br>(1151, 3146) | 7153<br>(3774, 12971) | 51545<br>(18000, 125363) | 58698<br>(25064, 134770) |
| Income classification |  |  |  |  |  |
| Low income | 1914<br>(1231, 2771) | 624<br>(413, 925) | 1648<br>(840, 2745) | 2435<br>(1339, 4394) | 4083<br>(2673, 6220) |
| Lower middle income | 2700<br>(1725, 4033) | 1431<br>(927, 2146) | 4762<br>(2473, 8613) | 15040<br>(8324, 26031) | 19803<br>(12533, 31084) |
| Upper middle income | 2312<br>(1065, 4437) | 1841<br>(1038, 3046) | 14136<br>(4012, 36858) | 96885<br>(39433, 203734) | 111021<br>(50911, 222528) |
| Time period |  |  |  |  |  |
| 2000-2009 | 865<br>(542, 1308) | 504<br>(318, 772) | 1592<br>(811, 3055) | 10203<br>(5078, 18940) | 11795<br>(6508, 20778) |
| 2010-2019 | 3710<br>(2356, 5554) | 2068<br>(1316, 3234) | 10751<br>(4932, 22492) | 61721<br>(27912, 122987) | 72472<br>(37745, 133518) |
| 2020-2023 | 2351<br>(1436, 3594) | 1324<br>(824, 2084) | 8203<br>(3404, 19386) | 42436<br>(19155, 83165) | 50640<br>(26167, 91817) |

The magnitude of averted utilization and costs increased over the study period, with 2020–2023 alone accounting for 34% of total averted utilization and 38% of total averted costs. Figure 1 presents time trends for these outcomes for countries with the highest burdens of each disease. In 2023, per 100 population, nearly 100 HIV-related outpatient visits were averted in Eswatini, and close to 80 bed-days were averted in Botswana and South Africa. Total averted costs per 100 population reached over US$7500 in Botswana. Similar trends were observed for TB and malaria, with the greatest reductions in PHC utilization and associated averted costs estimated for the final years of the time-series. Although the estimates of averted utilization and costs continued to rise over time, we saw a deceleration in the growth of these outcomes for most countries toward the end of the study period.

**Figure 1.**
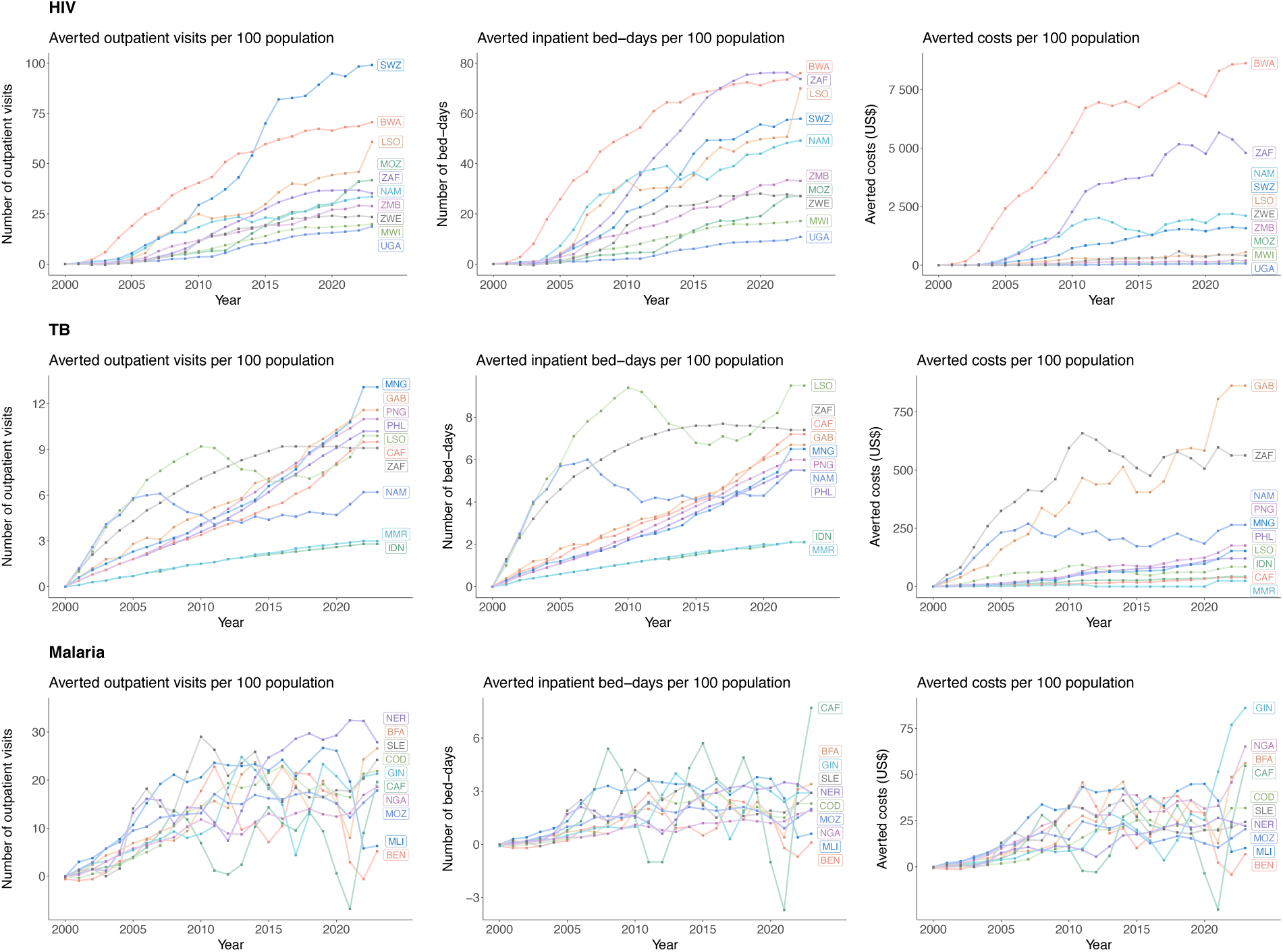
Averted primary health care utilization and associated averted costs due to scale-up of HIV, TB, and malaria services from 2000 to 2023, by year, country, and disease. Results shown for 10 modeled countries with the highest HIV prevalence, TB incidence rate, and malaria incidence rate in 2023.^34–36^

### Impact on health care capacity

We compared the estimates of averted inpatient bed-days to published estimates of hospital bed capacity for each country, to assess the magnitude of averted utilization relative to existing health system capacity (Figure 2). For 22 countries, the reduction in bed-days across all three diseases represented over 20% of hospital bed capacity over the study period. This impact was especially pronounced in the SSA region, where estimated reductions in hospital utilization were over 50% of national capacity for seven countries (Botswana, Lesotho, Ethiopia, Mozambique, Tanzania, Uganda, and South Africa). Across all SSA countries, the median reduction was 18·0% (interquartile range (IQR): 7·8–34·4) over the entire study period and 31·8% (IQR: 9·2–54·7) for 2023 (Table 2). Averted inpatient utilization represented 22·9% (IQR: 8·7–53·1) of hospital capacity in low-income countries for 2023, which was almost 4 times greater than the 6·3% (IQR: 1·2–22·3) in lower-middle-income countries and 13 times the percentages (1·7%, IQR: 0·5–3·5) in upper-middle-income countries.

**Figure 2.**
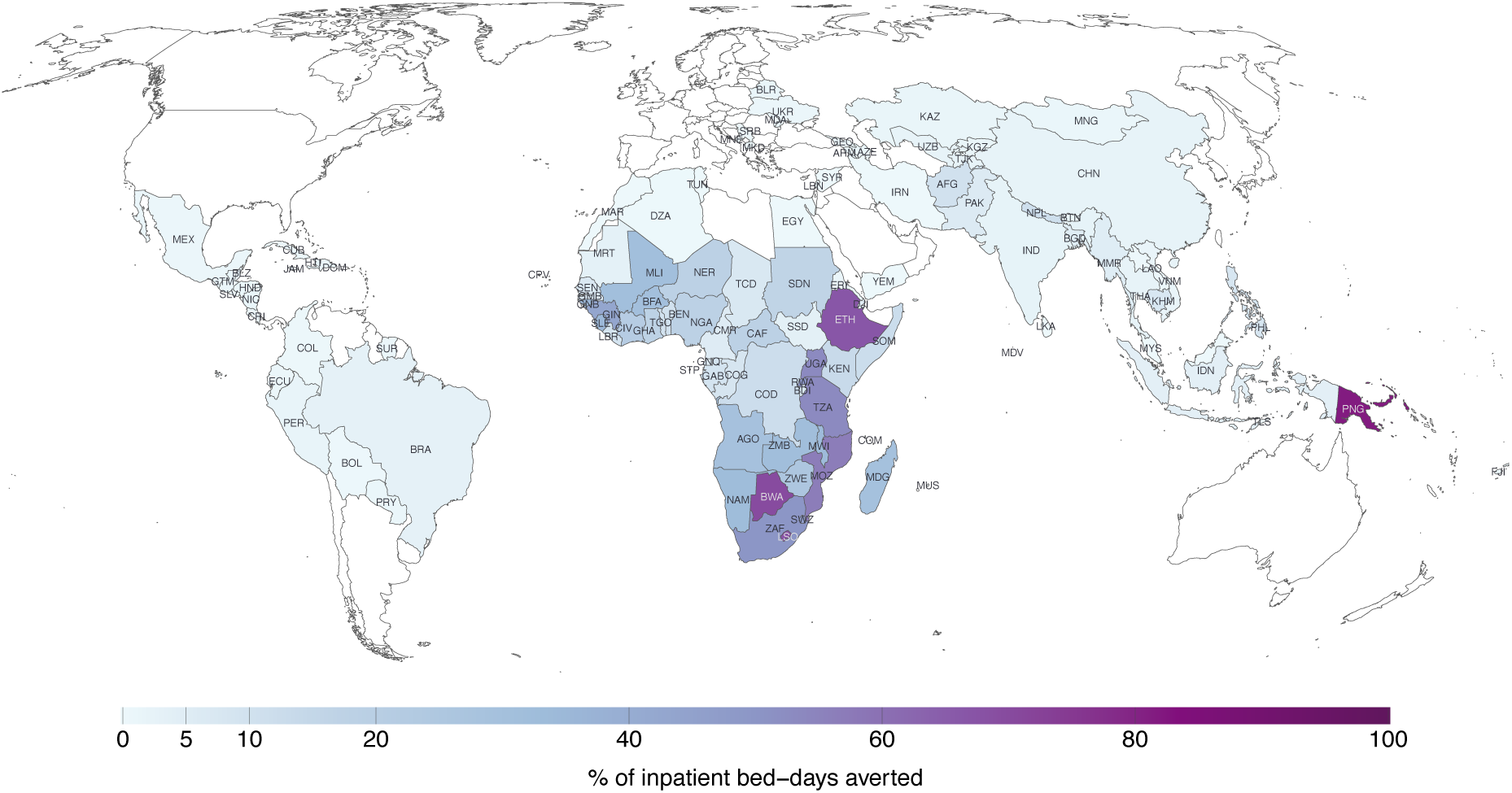
Averted inpatient bed-days due to scale-up of HIV, TB, and malaria services, as a percentage of national hospital bed capacity, 2000-23. Countries where averted inpatient utilization was over 50% of national hospital bed capacity: Papua New Guinea, Botswana, Lesotho, Ethiopia, Mozambique, Tanzania, Uganda, and South Africa.

**Table 2.** Estimates of averted inpatient utilization as a percentage of national hospital bed capacity and averted costs as a percentage of domestic government health expenditure, for actual scenario compared to constant coverage scenario, 2000-23. Point estimates represent median across modeled countries. Values in parentheses represent interquartile range (25^th^ and 75^th^ percentiles) of country-level values.

|  | Number of countries included in analysis | Averted inpatient bed-days (%) |  | Averted costs (%) |  |
| --- | --- | --- | --- | --- | --- |
|  |  | Whole period (2000-23) | Final year (2023) | Whole period (2000-23) | Final year (2023) |
| Total | 108 | 2.7<br>(0.6, 14.1) | 4.4<br>(1.1, 21.6) | 1.3<br>(0.1, 4.7) | 1.6<br>(0.2, 5.9) |
| Disease |  |  |  |  |  |
| HIV | 108 | 0.7<br>(0.2, 3.0) | 1.3<br>(0.4, 5.6) | 0.2<br>(0.05, 0.8) | 0.3<br>(0.07, 1.5) |
| TB | 51 | 4.9<br>(1.8, 9.6) | 7.2<br>(3.0, 16.1) | 1.5<br>(0.7, 2.0) | 2.5<br>(0.9, 4.0) |
| Malaria | 55 | 3.2<br>(0.2, 6.9) | 3.8<br>(0.1, 8.0) | 1.3<br>(0.1, 3.2) | 1.5<br>(0.07, 3.8) |
| World Bank region |  |  |  |  |  |
| East Asia and Pacific | 13 | 2.3<br>(0.4, 5.1) | 3.0<br>(0.7, 7.6) | 0.7<br>(0.3, 1.3) | 0.7<br>(0.3, 1.2) |
| Europe and Central Asia | 13 | 0.7<br>(0.2, 1.5) | 2.3<br>(0.7, 4.0) | 0.8<br>(0.2, 2.1) | 1.3<br>(0.3, 3.6) |
| Latin America and Caribbean | 18 | 0.9<br>(0.5, 1.1) | 1.5<br>(0.9, 2.3) | 0.1<br>(0.09, 0.2) | 0.2<br>(0.08, 0.3) |
| Middle East and North Africa | 9 | 0.1<br>(0.04, 0.14) | 0.2<br>(0.1, 0.5) | 0.03<br>(0.02, 0.05) | 0.08<br>(0.05, 0.1) |
| South Asia | 8 | 3.6<br>(0.1, 7.1) | 4.9<br>(0.3, 11.5) | 0.9<br>(0.04, 1.5) | 1.0<br>(0.06, 2.3) |
| Sub-Saharan Africa | 47 | 18.0<br>(7.8, 34.4) | 31.8<br>(9.2, 54.7) | 6.1<br>(2.2, 8.8) | 9.5<br>(2.7, 19.7) |
| Income classification |  |  |  |  |  |
| Low income | 25 | 16.3<br>(8.9, 33.7) | 22.9<br>(8.7, 53.1) | 4.4<br>(1.8, 8.8) | 5.1<br>(2.7, 14.0) |
| Lower middle income | 44 | 4.5<br>(0.8, 14.4) | 6.3<br>(1.2, 22.3) | 1.3<br>(0.3, 4.2) | 2.0<br>(0.4, 6.6) |
| Upper middle income | 39 | 0.9<br>(0.3, 1.8) | 1.7<br>(0.5, 3.5) | 0.2<br>(0.09, 0.8) | 0.3<br>(0.08, 1.2) |

### Impact on health care spending

To put the scale of financial benefits into perspective, we expressed the averted PHC costs as a fraction of published estimates of domestic government health expenditures. Across modeled countries, estimated averted costs represented a median of 1·3% (IQR: 0·1–4·7%) of total government health spending over the whole study period and 1·6% (IQR: 0·2–5·9%) for 2023 (Table 2). These percentages were higher for low-income countries (median in 2023: 5·1%, IQR: 2·7–14·0) compared to lower-middle-income countries (median in 2023: 2·0%, IQR: 0·4–6·6) and upper-middle-income countries (median in 2023: 0·3%, IQR: 0·08–1·2). Values for countries in the SSA region clustered around the 10% reference line, with averted PHC costs for 2023 surpassing 20% of government health spending in 12 out of 47 countries in this region (Figure 3).

**Figure 3.**
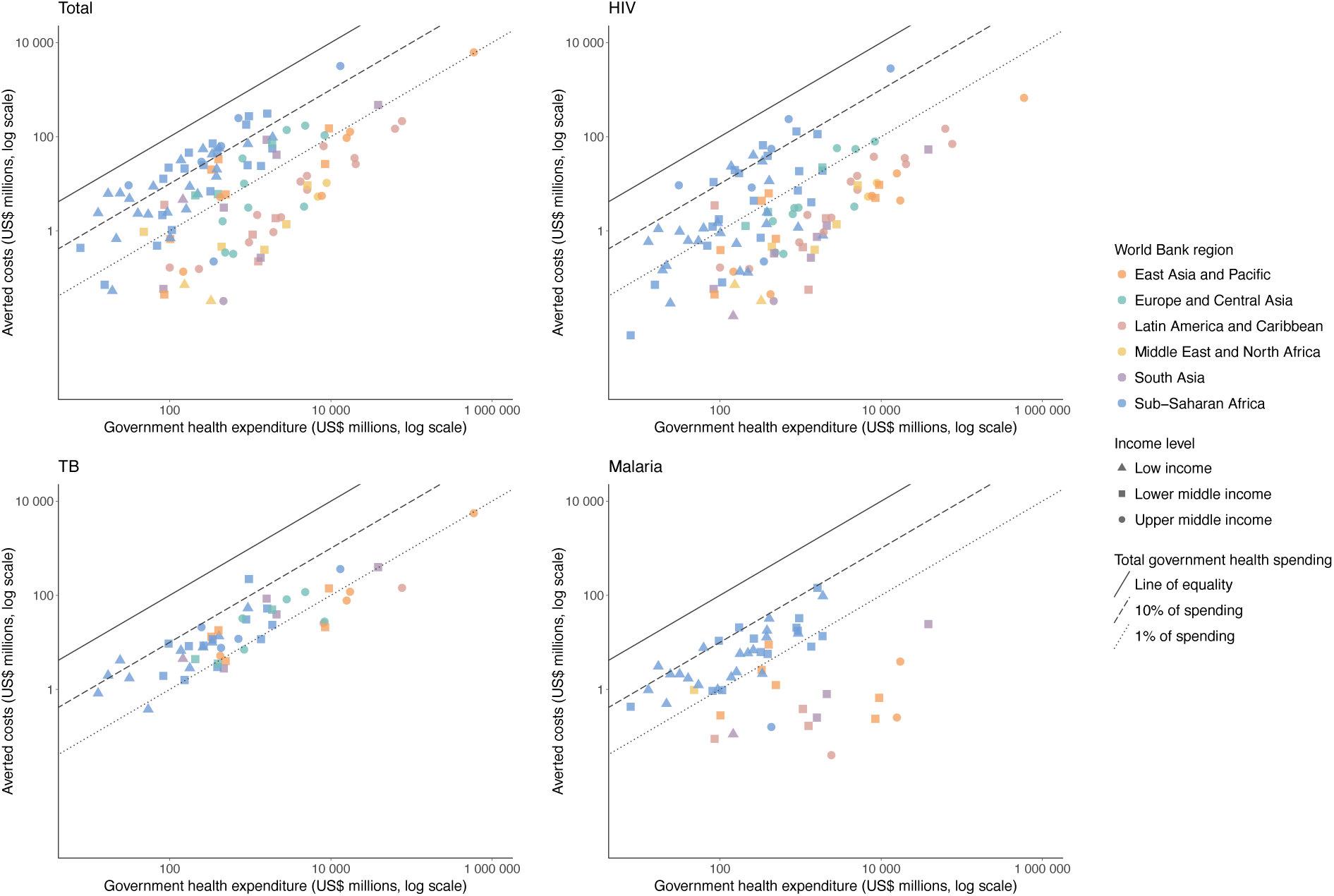
Averted costs due to scale-up of HIV, TB, and malaria services compared to domestic government health expenditures by country in 2023. Plotted values represent nominal U.S. dollars.

### Sensitivity analyses

Tables S-3 and S-4 report results for the *actual scenario* compared to the *no coverage scenario*, which assumed no HTM services after year 2000. As expected, these results were larger in magnitude than those from the main analysis. An estimated 18·3 (10·3–29·7) billion outpatient visits and 10·1 (6·2–15·5) billion inpatient bed-days were averted, 2·6 times greater than the values reported in the main analysis. Averted costs totaled US$358 (184–657) billion over the study period, 2·7 times higher than estimated in the main analysis.

Figure S-2 presents PRCCs describing the relationship between individual parameters and total averted costs estimated for each disease. For HIV and TB, the most influential parameters were the duration of hospitalization and the unit cost per hospital bed-day. For malaria, the most influential determinants were the difference in symptomatic malaria cases between scenarios, the number of outpatient visits per malaria case, and the unit cost per outpatient visit.

Table S-5 compares the results of our main analysis to two sensitivity analyses that allowed PHC unit costs to vary to reflect economies or diseconomies of scale. When we allowed for a negative relationship between PHC service volume and unit costs, overall averted cost estimates were 20% lower than in the main analysis. When we assumed that increased utilization would exceed capacity constraints and require additional investments, averted costs were 25% greater than in the main analysis.

## Discussion

We investigated how investments to scale-up of HIV, TB, and malaria services have influenced primary health care in 108 LMICs, specifically through changes in health care utilization among persons with untreated HIV, TB, or malaria who are unable to access effective disease-specific care. We estimated substantial reductions in PHC utilization among these persons during 2000–2023, with 6·9 billion outpatient visits and 3·9 billion inpatient bed-days averted over the period, as compared to a counterfactual scenario constraining HTM service coverage at year 2000 levels. This reduction in utilization represents US$135 billion in averted costs. Additionally, for several countries, particularly those with high disease burden and limited health system capacity, the reductions in PHC utilization and costs were large relative to national hospital capacity and government health spending.

Several previous studies have suggested pathways through which disease-specific investments can enhance health service delivery, improve medical supply chains, reduce the burden of related conditions, and facilitate human resources for health in LMICs.^22–25^ The results of this study demonstrate another mechanism through which these investments can contribute to health system strengthening, not usually quantified in other analyses. This work adds to the body of literature documenting the complex consequences of global health initiatives to address specific diseases in resource-limited settings.^26–29^ Under the *constant coverage* counterfactual scenario, low HTM service coverage allowed for continued disease evolution and spread, and any reductions in disease burden achieved pre-2000 plateaued and reversed. As a consequence, low HTM service coverage was projected to produce an increasing number of individuals with symptomatic HIV, TB, or malaria without effective disease-specific treatment, who would seek primary care when symptoms emerged, and potentially require hospitalization with the development of advanced conditions. In the sensitivity analysis where we examined the *no coverage* counterfactual scenario, each disease was projected to expand unchecked, leading to larger numbers of symptomatic individuals seeking primary health care in subsequent years. The projected surge in demand for health services under the counterfactual scenarios illustrates the impact that the scale-up of HTM services has had in averting preventable PHC utilization and costs.

Overall, our estimates showed progressive growth in the magnitude of averted PHC utilization and costs over the study period, as HTM services were scaled up and the compounding benefits of transmission reductions were realized. An exception to this pattern was malaria, for which we identified fluctuations in the time trends of estimated outcomes for some countries and years, especially around 2020. These were likely attributable to resurgences of malaria incidence driven by changes in funding mechanisms, COVID-19 pandemic-related disruptions, and multi-year cycles for mass distributions of insecticide-treated bed nets.^30–32^ This finding reveals how interruptions in intervention access can have secondary consequences on other health care, adding resource and financial burdens to the already strained primary health care systems.

In sensitivity analyses, we explored how changes in the cost functions used to calculate PHC costs would affect cost results. The main analysis assumed that PHC unit costs would not change with different utilization levels, such that any change in utilization would produce a proportional change in total costs. When we allowed for unit costs to decline with increasing utilization due to economies of scale, the averted costs were 20% smaller. When we allowed unit costs to rise with increasing utilization due to the costs of expanding capacity, the averted costs were 25% larger. The balance of these two mechanisms is unknown and will likely differ between countries and according to the magnitude of additional utilization. Nevertheless, it is notable that the size of estimated averted costs was substantial under each of the approaches we explored.

This study has several limitations. First, we did not consider reductions in health care utilization resulting from premature mortality. It is possible that the estimated reductions in PHC utilization would be offset by continued utilization by individuals whose deaths were averted by HTM services. Second, there is limited evidence on health care utilization rates for individuals with untreated HIV, TB, or malaria, increasing the uncertainty of our estimates. Third, our analysis did not consider changes in PHC services beyond utilization. Changes in utilization could also affect service quality, timeliness, and patient satisfaction, but these outcomes were not considered in this study. We also did not estimate resource needs and utilization within HTM programs themselves, as these have been estimated by other analyses.^33^ Fourth, although we calculated outcomes using country-level epidemiological data, several analytic inputs were only available as global averages or were translated to different country settings using regression adjustments. For this reason, outcomes for individual countries should be interpreted with caution. Lastly, some population groups can face systemic access barriers, limiting utilization of both HTM and PHC services. We did not investigate the implications of these correlated patterns of disadvantage. Given these limitations, further research is needed to illuminate the societal and economic impacts of investments to combat major infectious diseases across different LMIC settings, including how supply-side factors play a role in mediating these outcomes.

This analysis provides evidence on how investments to scale up HIV, TB, and malaria services over the start of the 21^st^ century have reduced the burden placed by these diseases on routine health systems. Our findings underscore the positive contribution that sustained support by international donors and domestic governments for HIV, TB, and malaria elimination can make to the broader agenda of promoting health system resilience and efficiency. The results highlight the importance of recognizing not only the direct health and economic gains from these investments but also the indirect benefits for primary health systems. These indirect impacts should be explicitly considered when evaluating the full value of global health investments.

## Data Availability

All data produced in the present study are available upon reasonable request to the authors.

## Acknowledgements

The authors thank Mehran Hosseini, Richard Grahn, Mikaela Smit, and Johannes Hunger for feedback on early versions of this analysis.

## Author Contributions

NAM conceptualized the study. NAM and JSS designed the analysis with input from all authors. JS, CP, and PW conducted epidemiological modeling. JSS conducted the analysis, produced the results, and drafted the manuscript. NAM and JSS revised the manuscript. All authors reviewed and edited the final version of the manuscript.

