## Supplementary Appendix for "The benefits of investments to combat HIV, tuberculosis and malaria for primary health care, 2000–2023: an economic modeling analysis"

**Supplementary methods**

*TB emulator for additional TB-burden countries*

For each scenario, we extracted estimates of epidemiological outcomes for 29 countries from the results of the *Global Portfolio TB Model,*[1] covering 81% of global TB incidence. To increase the fraction of global TB burden covered by our analysis, we developed a regression-based emulator using the modeled results for 29 countries. The emulator was constructed as two generalized additive models (GAMs), predicting the TB incidence rate and TB treatment coverage, respectively. For the TB incidence rate, we developed a GAM model using a log link function and a Gaussian error distribution. Predictors included smooth terms for the log-transformed TB incidence rate (derived from the WHO burden estimates),[2] the percentage of incident TB cases that are HIV-positive,[2] and calendar year. The number of smooth terms for modelled predictors was chosen to minimize root mean square prediction error based on leave-one-out cross-validation. For the *actual scenario*, the annual number of new TB cases was calculated by multiplying the TB incidence rate (as predicted by the regression model) by annual population. For the *constant coverage scenario* and *the no coverage scenario*, we refit the emulation model using the ratio of annual TB cases in each counterfactual scenario relative to the number of annual TB cases in the *actual scenario* as the predicted variable. We set year 2000 as the base year, for which outcomes were assumed to match those of the *actual scenario*. The final estimates of new TB cases in counterfactual scenarios were then imputed as the number of new TB cases in the *actual scenario* multiplied by the ratios estimated from the regression model.

To model treatment coverage, we fit a GAM model with a logit link function and a Gaussian error distribution. The model included smooth terms for logit-transformed treatment coverage,[2] the percentage of incident TB cases that are HIV-positive,[2] and calendar year. The predicted variable for the *actual scenario* was the TB treatment coverage. The predicted variable for the *constant coverage scenario* was the ratio of TB treatment coverage in this scenario compared to the *actual scenario*. We set year 2000 as the base year, and calculated treatment coverage in the *constant coverage scenario* as treatment coverage for the *actual scenario* multiplied by the ratios estimated from the regression model. Treatment coverage for the *no coverage scenario* was set to zero over the study period.

Using this emulator, we predicted epidemiological results for each scenario for an additional set of 22 countries not included in the original modeled results. These countries were chosen based on their inclusion in WHO lists of high-burden countries (for TB, TB-HIV, or MDR-TB)[3] and covered 11% of global TB incidence.

*Sensitivity analyses: Changes in unit costs*

We conducted two additional sensitivity analyses in which we explored non-linear cost functions describing how total PHC costs would change under different levels of utilization. In the first of these sensitivity analyses, we adopted a cost model allowing for economies of scale, in which the cost per outpatient visit and per inpatient bed-day was assumed to decrease as utilization increases. This was operationalized as an elasticity relationship, in which *ln(unit cost) ~ ln(utilization)*b*. In this relationship, a negative value of *b* allows for reductions in the unit cost for higher levels of utilization. Estimates of this parameter were based on published econometric analyses.[4,5]

In the second sensitivity analysis, we modified the cost function from the main analysis to explore the additional costs of relaxing capacity constraints with higher levels of PHC utilization, as might be required for infrastructure investments and staff training. To implement this analysis, we extracted data on current levels of facility capacity utilization collected as part of a multi-country cost study.[6] We assumed that any increase in utilization would affect all facilities proportionally, and recalculated capacity utilization results for higher levels of utilization. For facilities in which these revised capacity utilization estimates exceeded 100%, we assumed the excess utilization would require additional fixed costs to expand capacity. This was calculated as a 62% mark-up on the original unit cost, applied to utilization above 100% of the original capacity.[6]

For both of these sensitivity analyses, we applied the adjusted cost functions to the changes in PHC utilization estimated in our main analysis, for each country and year. The resulting averted costs were compared to the averted cost estimates from our main analysis to evaluate the influence of these alternative assumptions.

**Figure S-1a. Flow chart of outcomes for HIV.**

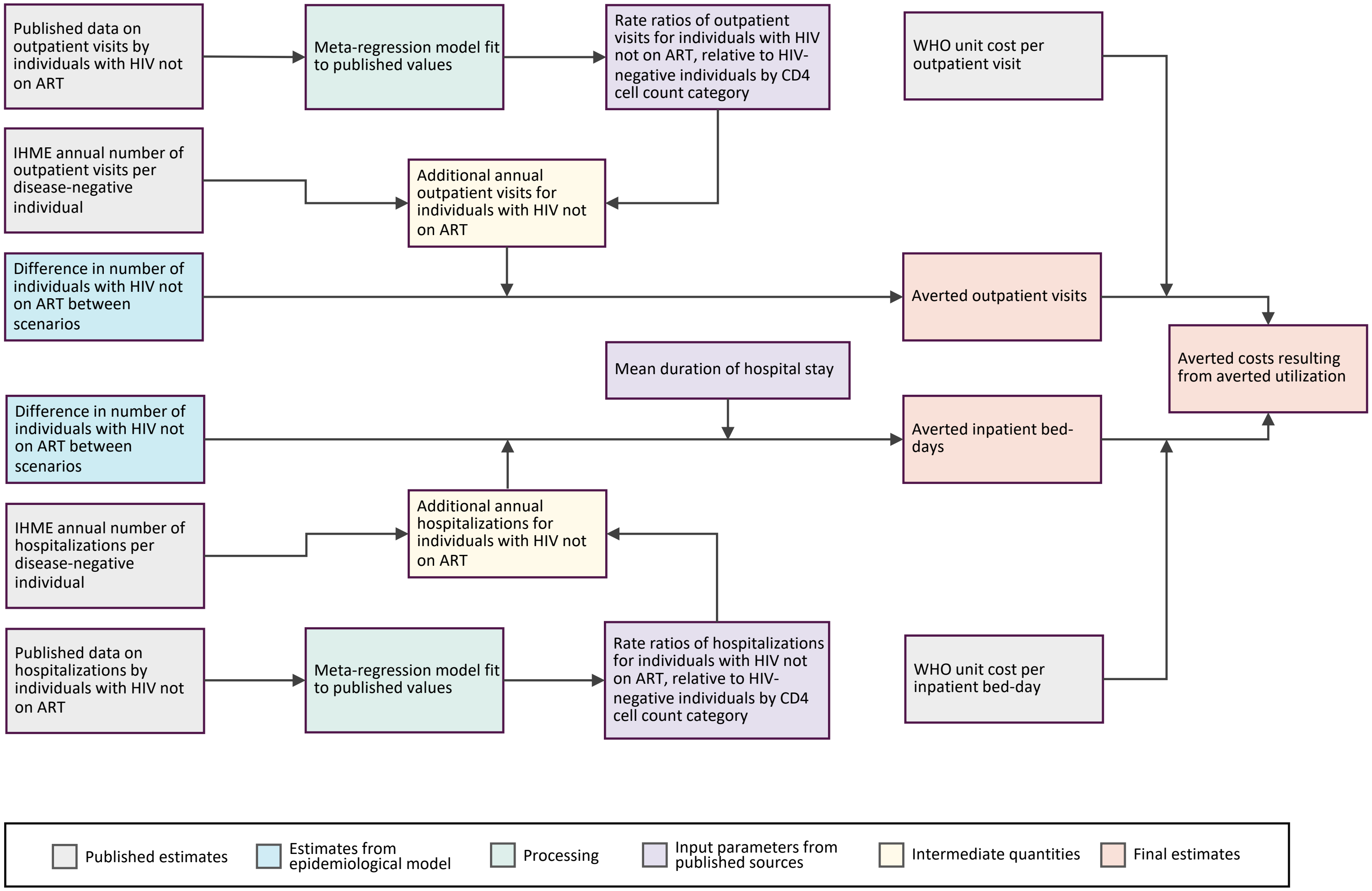

**Figure S-1b. Flow chart of outcomes for TB.**

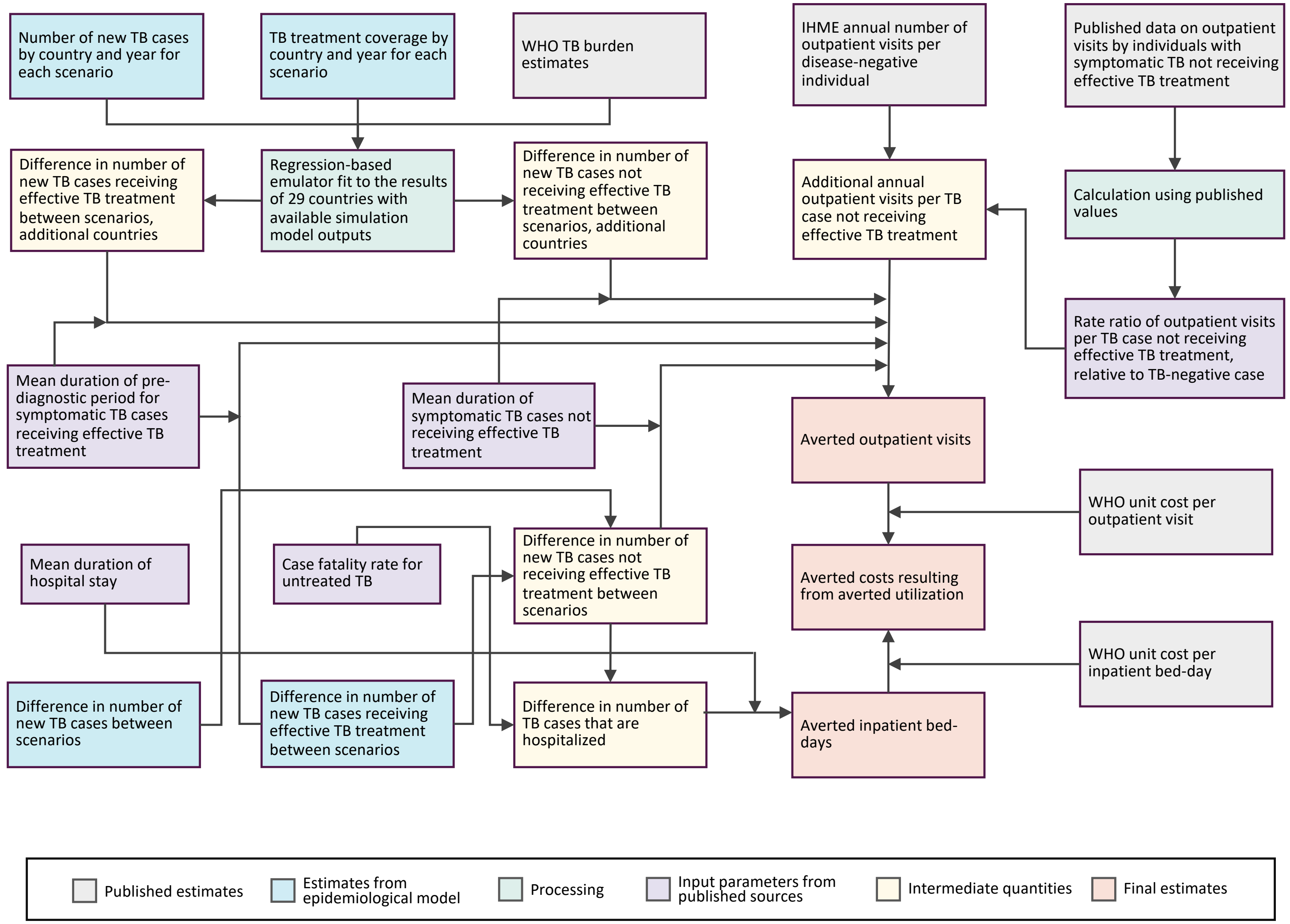

**Figure S-1c. Flow chart of outcomes for malaria.**

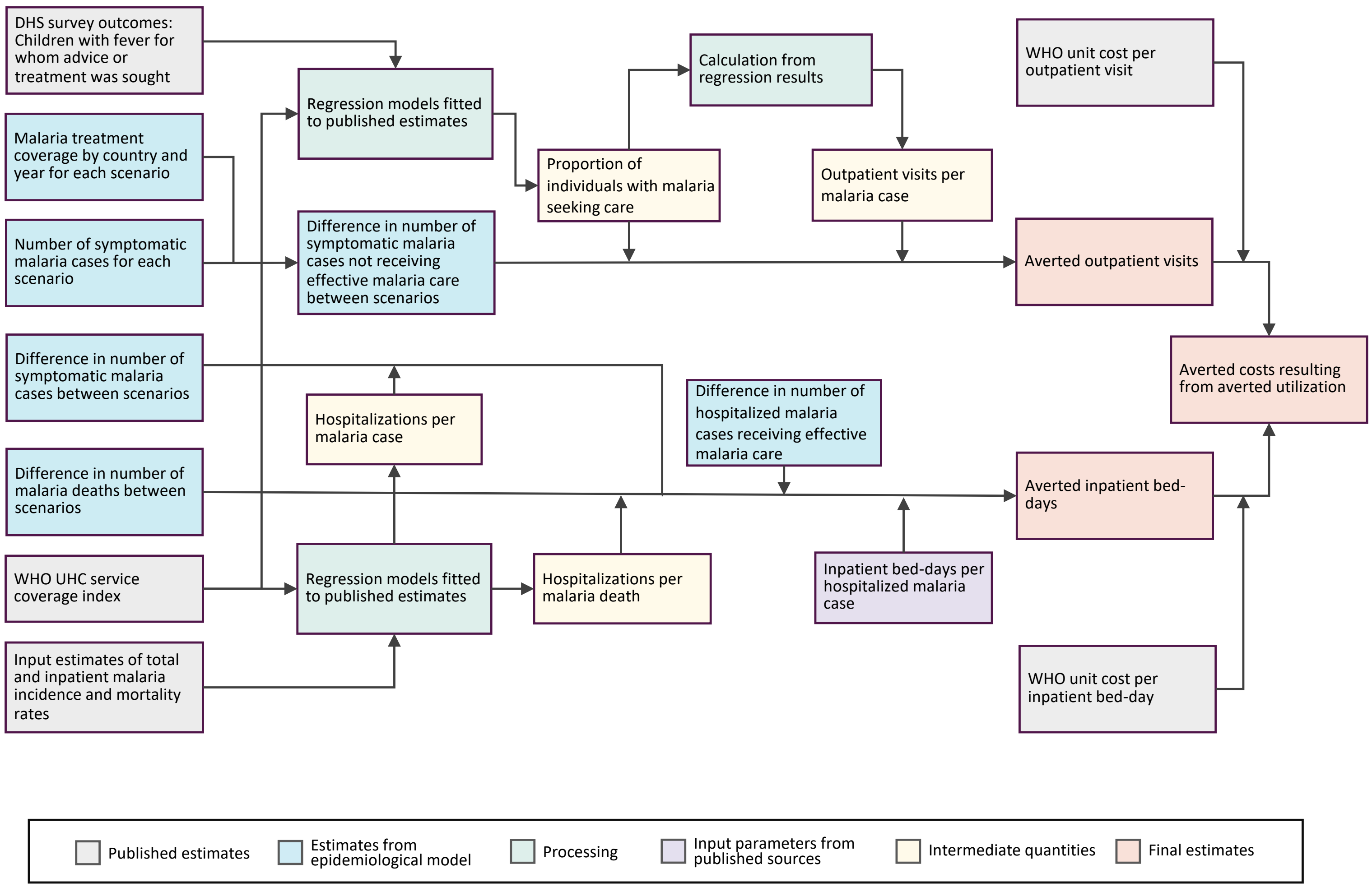

**Table S-1a. Summary of HIV input parameters.**

| Input parameter | Mean value (95% uncertainty interval) ‡ | Distribution † | Reference |
|---|---|---|---|
| Number of individuals with HIV not on ART | Stratified by country, year, age group, CD4 cell count category, and scenario | Gamma | Epidemiological models ʔ |
| Population | Stratified by country and year | Fixed value | |
| Annual number of outpatient visits per disease-negative individual | Stratified by country, year, and age group | Gamma | IHME healthcare utilization estimates[7] |
| Annual number of hospitalizations per disease-negative individual | Stratified by country, year, and age group | Gamma | |
| Rate ratio of outpatient visits for individuals with HIV not on ART, relative to HIV-negative individuals | | Gamma | Estimated from meta-regression model fit to published values[8-12] |
| CD4 count 0-49 cells/mm$^3$ | 4.4 (1.9, 9.0) | | |
| CD4 count 50-99 cells/mm$^3$ | 4.0 (1.8, 7.9) | | |
| CD4 count 100-199 cells/mm$^3$ | 3.5 (1.7, 6.4) | | |
| CD4 count 200-249 cells/mm$^3$ | 3.0 (1.6, 5.3) | | |
| CD4 count 250-349 cells/mm$^3$ | 2.7 (1.5, 4.3) | | |
| CD4 count 350-499 cells/mm$^3$ | 2.1 (1.4, 3.1) | | |
| CD4 count ≥ 500 cells/mm$^3$ | 1.5 (1.2, 1.8) | | |
| Rate ratio of hospitalization for individuals with HIV not on ART, relative to HIV-negative individuals | | Gamma | Estimated from meta-regression model fit to published values[8-11,13] |
| CD4 counts 0-49 cells/mm$^3$ | 42.1 (13.9, 99.4) | | |
| CD4 counts 50-99 cells/mm$^3$ | 33.3 (11.8, 75.2) | | |
| CD4 counts 100-199 cells/mm$^3$ | 23.5 (9.3, 49.5) | | |
| CD4 counts 200-249 cells/mm$^3$ | 16.6 (7.3, 32.6) | | |
| CD4 counts 250-349 cells/mm$^3$ | 11.8 (5.8, 21.5) | | |
| CD4 counts 350-499 cells/mm$^3$ | 6.7 (3.9, 10.7) | | |
| CD4 counts ≥ 500 cells/mm$^3$ | 2.7 (2.0, 3.5) | | |
| Mean duration of hospital stay, days | 6.6 (4.0, 13.0) | Gamma | Published estimates[12,13] |
| Cost per outpatient visit § | Stratified by country and year | Gamma | WHO-CHOICE estimates[4] |
| Cost per inpatient bed-day § | Stratified by country and year | Gamma | WHO-CHOICE estimates[4] |

‡ We assumed a 95% interval equivalent to +/- 50% of the point estimate for parameters for which measures of uncertainty were not available.
† To create prior distributions for probabilistic sensitivity analysis, we specified distributions matching the mean value and interval width for each input parameter.
ʔ Epidemiological outcomes for each disease and scenario were obtained by country and year from models employed for the Global Fund 8th replenishment investment case.[14] These models included the Goals Model for HIV, the Global Portfolio TB Model for TB, and the *malariasimulation* model for malaria.[1,15,16]
§ Same unit cost (cost per outpatient visit, cost per inpatient bed-day) values were applied to all three diseases.

**Table S-1b. Summary of TB model input parameters.**

| Input parameter | Mean value (95% uncertainty interval) [‡] | Distribution [†] | Reference |
|---|---|---|---|
| Number of new TB cases | Stratified by country, year, and scenario | Gamma | Epidemiological models [ʔ] |
| Number of new TB cases receiving effective TB treatment | Stratified by country, year, and scenario | Gamma | |
| Number of new TB cases, additional countries | Stratified by country, year, and scenario | Multivariate normal for logged outcome | Imputed using model emulator [Ψ] |
| Treatment coverage, additional countries | Stratified by country, year, and scenario | Multivariate normal for log-odds of outcome | |
| Mean outpatient visits per disease-negative individual | Stratified by country and year | Gamma | IHME healthcare utilization estimates[7] |
| Mean duration of pre-diagnostic period for symptomatic TB cases receiving effective TB treatment, months | 1.0 (0.5, 1.5) | Gamma | Published estimates[17,18] |
| Rate ratio of outpatient visits per TB case not receiving effective TB treatment, relative to TB-negative case | 6.23 (4.49, 11.47) | Gamma | Published estimates[19-27] |
| Mean duration of symptomatic TB cases not receiving effective TB treatment, months | 6.0 (3.0, 9.0) | Gamma | Published estimates of untreated disease duration,[28,29] adjusted for symptom prevalence[30] [δ] |
| Case fatality rate for untreated TB | 0.43 (0.28, 0.53) | Beta | WHO Global TB Program Methods Supplement[31] |
| Mean duration of hospital stay, days | 14.0 (7.0, 21.0) | Gamma | WHO TB patient cost surveys 2015-2021[32] |
| Cost per outpatient visit [§] | Stratified by country and year | Gamma | WHO-CHOICE estimates[4] |
| Cost per inpatient bed-day [§] | Stratified by country and year | Gamma | WHO-CHOICE estimates[4] |

‡ We assumed a 95% interval equivalent to +/- 50% of the point estimate for parameters where measures of uncertainty were not available.
† To create prior distributions for probabilistic sensitivity analysis, we specified distributions matching the mean value and interval width for each input parameter.
ʔ Epidemiological outcomes for each disease and scenario were obtained by country and year from models employed for the Global Fund 8th replenishment investment case.[14] These models included the Goals Model for HIV, the Global Portfolio TB Model for TB, and the *malariasimulation* model for malaria.[1,15,16]
Ψ For 22 countries, values were estimated using a regression-based emulator fit to the results of 29 countries with available simulation model outputs, using WHO burden estimates.[2]
δ There is substantial uncertainty about the duration of untreated TB (estimates range from 1.6 to 5.4 years) and the prevalence of symptoms (estimates range from 36 to 80%). Our estimate (mean 6 months) represents a conservative value based on available evidence.
§ Same unit cost (cost per outpatient visit, cost per inpatient bed-day) values were applied to all three diseases.

**Table S-1c. Summary of malaria model input parameters.**

| Input parameter | Mean value (95% uncertainty interval) ‡ | Distribution † | Reference |
|---|---|---|---|
| Number of symptomatic malaria cases | Stratified by country, year, and scenario | Gamma | Epidemiological models ˀ |
| Treatment coverage | Stratified by country, year, and scenario | Fixed value | |
| Number of malaria deaths | Stratified by country, year and scenario | Gamma | |
| Number of hospitalized malaria cases receiving effective malaria care | Stratified by country, year and scenario | Fixed value | |
| Outpatient visits per malaria case | 1.0 (0.5, 1.5) | Gamma | Imputed from published estimates[33] |
| Proportion of individuals with malaria seeking care | Stratified by country and year | Multivariate normal for log-odds of outcome | Estimated from regression model fit to published values[33] ø |
| Hospitalizations per malaria case | Stratified by country | Multivariate normal for logged outcome | Estimated from regression model fit to published values[34,35] ^ |
| Hospitalizations per malaria death | Stratified by country | Multivariate normal for logged of outcome | Estimated from regression model fit to published values[34,35] ^ |
| Inpatient bed-days per hospitalized malaria case | 3.0 (1.5, 4.5) | Gamma | Published estimate[36] |
| Cost per outpatient visit § | Stratified by country and year | Gamma | WHO-CHOICE estimates[4] |
| Cost per inpatient bed-day § | Stratified by country and year | Gamma | WHO-CHOICE estimates[4] |

‡ We assumed a 95% interval equivalent to +/- 50% of the point estimate for parameters where measures of uncertainty were not available.
† To create prior distributions for probabilistic sensitivity analysis, we specified distributions matching the mean value and interval width for each input parameter.
ˀ Epidemiological outcomes for each disease and scenario were obtained by country and year from models employed for the Global Fund 8th replenishment investment case.[14] These models included the Goals Model for HIV, the Global Portfolio TB Model for TB, and the *malariasimulation* model for malaria.[1,15,16]
ø We fit a generalized linear mixed model with a logit link function and a Gaussian error distribution to calculate the proportion of malaria individuals seeking care, for which we included random effects for country, a linear term for time (centered on year 2000), and a covariate for the universal health coverage index.[35]
^ We developed respective log-linear regression models to estimate the number of hospitalizations per malaria case and per malaria death as a function of the universal health coverage index.[35]
§ Same unit cost (cost per outpatient visit, cost per inpatient bed-day) values were applied to all three diseases.

**Table S-2a. Cumulative averted primary health care utilization and associated averted costs due to scale-up HIV services, for actual scenario compared to constant coverage scenario, 2000-23.**

| | Averted utilization, millions | | Averted costs (US$), millions | | |
|---|---|---|---|---|---|
| | Outpatient visits | Inpatient bed-days | Outpatient | Inpatient | Total |
| Total | 1188 (757, 1734) | 1352 (516, 2710) | 4041 (1927, 7677) | 50121 (14679, 124034) | 54161 (17960, 129606) |
| World Bank region | | | | | |
| East Asia and Pacific | 111 (67, 163) | 113 (40, 222) | 558 (153, 1428) | 5349 (1246,13268) | 5907 (1724, 13829) |
| Europe and Central Asia | 27 (17, 40) | 33 (13, 64) | 129 (53, 256) | 1260 (413, 2788) | 1389 (540, 2956) |
| Latin America and Caribbean | 89 (56, 126) | 73 (28, 149) | 453 (188, 903) | 2743 (885, 6327) | 3195 (1267, 6826) |
| Middle East and North Africa | 4.0 (2.5, 5.8) | 3.4 (1.3, 6.8) | 25 (10, 52) | 165 (54, 379) | 190 (73, 413) |
| South Asia | 54 (28, 88) | 47 (16, 101) | 91 (12, 257) | 482 (109, 1334) | 573 (165, 1408) |
| Sub-Saharan Africa | 903 (577, 1327) | 1083 (412, 2175) | 2786 (1030, 6562) | 40122 (9543, 111749) | 42908 (12844, 113221) |
| Income classification | | | | | |
| Low income | 273 (169, 402) | 183 (71, 364) | 215 (94, 423) | 619 (211, 1353) | 833 (391, 1612) |
| Lower middle income | 460 (286, 674) | 424 (166, 874) | 741 (392, 1251) | 3778 (1432, 7871) | 4519 (2177, 8800) |
| Upper middle income | 455 (282, 690) | 744 (256, 1627) | 3085 (1169, 6726) | 45724 (11949, 119343) | 48809 (14829, 121741) |
| Time period | | | | | |
| 2000-2009 | 79 (45, 124) | 99 (36, 202) | 222 (100, 411) | 3081 (836, 8297) | 3303 (1064, 8567) |
| 2010-2019 | 627 (401, 923) | 735 (274, 1470) | 2192 (1000, 4359) | 28475 (7987, 71622) | 30668 (9873, 74601) |
| 2020-2023 | 482 (310, 692) | 518 (203, 1018) | 1626 (791, 3027) | 18565 (5551, 45056) | 20191 (7064, 46936) |

Values in parentheses represent 95% uncertainty intervals. Cost values represent nominal U.S. dollars. Actual scenario represents the observed scale-up of HIV services over the study period. Constant coverage scenario represents a counterfactual with coverage of HIV services held constant at the year 2000 levels for each country over the study period.

**Table S-2b. Cumulative averted primary health care utilization and associated averted costs due to scale-up TB services, for actual scenario compared to constant coverage scenario, 2000-23.**

| | Averted utilization, millions | | Averted costs (US$), millions | | |
|---|---|---|---|---|---|
| | Outpatient visits | Inpatient bed-days | Outpatient | Inpatient | Total |
| Total | 3303 (1127, 6816) | 2150 (1164, 3579) | 13447 (3357, 36657) | 61431 (24074, 131111) | 74878 (35791, 148883) |
| World Bank region | | | | | |
| East Asia and Pacific | 1687 (565, 3621) | 1009 (512, 1754) | 9744 (1342, 31835) | 43538 (13545, 104946) | 53282 (19148, 118791) |
| Europe and Central Asia | 180 (61, 372) | 62 (33, 104) | 780 (209, 1985) | 2158 (926, 4208) | 2938 (1466, 5352) |
| Latin America and Caribbean | 155 (53, 337) | 83 (40, 147) | 360 (55, 1101) | 1610 (488, 3870) | 1970 (705, 4236) |
| Middle East and North Africa | - | - | - | - | - |
| South Asia | 622 (205, 1237) | 492 (267, 823) | 978 (175, 2705) | 4846 (1601, 10821) | 5824 (2242, 12171) |
| Sub-Saharan Africa | 659 (222, 1334) | 505 (276, 843) | 1585 (439, 3902) | 9280 (3712, 18979) | 10865 (5113, 21027) |
| Income classification | | | | | |
| Low income | 316 (106, 640) | 234 (128, 390) | 237 (67, 608) | 728 (322, 1428) | 965 (506, 1797) |
| Lower middle income | 1136 (375, 2279) | 821 (446, 1370) | 2173 (661, 5031) | 9575 (4169, 18406) | 11748 (5911, 20949) |
| Upper middle income | 1852 (621, 3982) | 1095 (553, 1908) | 11038 (2078, 33578) | 51127 (18118, 117237) | 62166 (26432, 131016) |
| Time period | | | | | |
| 2000-2009 | 393 (132, 813) | 343 (184, 577) | 964 (279, 2344) | 6758 (2821, 13604) | 7722 (3767, 14564) |
| 2010-2019 | 1679 (575, 3507) | 1089 (589, 1816) | 6788 (1755, 18513) | 31522 (12560, 67730) | 38311 (18414, 75440) |
| 2020-2023 | 1232 (415, 2515) | 719 (380, 1206) | 5695 (1303, 16597) | 23151 (8729, 50518) | 28846 (13138, 58750) |

Values in parentheses represent 95% uncertainty intervals. Cost values represent nominal U.S. dollars. Actual scenario represents the observed scale-up of TB services over the study period. Constant coverage scenario represents a counterfactual with coverage of TB services held constant at the year 2000 levels for each country over the study period.

**Table S-2c. Cumulative averted primary health care utilization and associated averted costs due to scale-up malaria services, for actual scenario compared to constant coverage scenario, 2000-23.**

| | Averted utilization, millions | | Averted costs (US$), millions | | |
|---|---|---|---|---|---|
| | Outpatient visits | Inpatient bed-days | Outpatient | Inpatient | Total |
| Total | 2435 (1371, 3872) | 395 (184, 767) | 3058 (1455, 5423) | 2808 (1186, 6041) | 5866 (3344, 9539) |
| World Bank region | | | | | |
| East Asia and Pacific | 39 (20, 65) | 8.1 (3.5, 16.8) | 90 (24, 235) | 97 (38, 213) | 187 (86, 358) |
| Europe and Central Asia | - | - | - | - | - |
| Latin America and Caribbean | 1.1 (0.6, 1.9) | 0.4 (0.1, 1.4) | 2.7 (0.9, 5.9) | 6.2 (0.9, 23.1) | 8.9 (2.6, 26.3) |
| Middle East and North Africa | 1.3 (0.6, 2.3) | 0.3 (0.1, 1.0) | 3.0 (0.3, 9.1) | 4.3 (0.8, 15.4) | 7.3 (1.9, 19.8) |
| South Asia | 115 (54, 210) | 56 (10, 165) | 181 (25, 511) | 558 (65, 2066) | 739 (159, 2237) |
| Sub-Saharan Africa | 2279 (1279, 3618) | 330 (159, 591) | 2782 (1325, 5067) | 2143 (965, 4369) | 4925 (2803, 7880) |
| Income classification | | | | | |
| Low income | 1326 (718, 2097) | 207 (94, 387) | 1197 (555, 2221) | 1088 (404, 2777) | 2285 (1212, 4232) |
| Lower middle income | 1104 (601, 1779) | 186 (84, 384) | 1848 (750, 3823) | 1687 (685, 3702) | 3535 (1912, 6115) |
| Upper middle income | 5.1 (2.5, 9.2) | 2.1 (0.6, 4.7) | 13 (2, 39) | 33 (5, 97) | 46 (14, 116) |
| Time period | | | | | |
| 2000-2009 | 393 (221, 619) | 62 (30, 124) | 406 (189, 755) | 364 (152, 813) | 770 (439, 1309) |
| 2010-2019 | 1405 (799, 2225) | 245 (112, 475) | 1771 (843, 3121) | 1723 (719, 3813) | 3494 (2002, 5699) |
| 2020-2023 | 637 (355, 1010) | 88 (40, 164) | 882 (413, 1602) | 721 (303, 1573) | 1603 (896, 2551) |

Values in parentheses represent 95% uncertainty intervals. Cost values represent nominal U.S. dollars. Actual scenario represents the observed scale-up of malaria services over the study period. Constant coverage scenario represents a counterfactual with coverage of malaria services held constant at the year 2000 levels for each country over the study period.

**Table S-3. Sensitivity analysis – Cumulative averted primary health care utilization and associated averted costs due to scale-up HIV, TB, and malaria services, for actual scenario compared to no coverage scenario, 2000-23.**

| | Averted utilization, millions | | Averted costs (US$), millions | | |
|---|---|---|---|---|---|
| | Outpatient visits | Inpatient bed-days | Outpatient | Inpatient | Total |
| Total | 18280 (10296, 29656) | 10126 (6168, 15496) | 58175 (22374, 142411) | 300104 (138028, 582074) | 358279 (184158, 657314) |
| Disease | | | | | |
| HIV | 1580 (998, 2310) | 2032 (772, 4178) | 6795 (2509, 16027) | 93789 (22604, 252164) | 100583 (29596, 262865) |
| TB | 11159 (3754, 22597) | 7313 (3920, 12333) | 44254 (11065, 128923) | 200448 (77535, 433993) | 244701 (110162, 487506) |
| Malaria | 5541 (3077, 8777) | 781 (380, 1511) | 7126 (3343, 13045) | 5868 (2458, 13002) | 12994 (7539, 21852) |
| World Bank region | | | | | |
| East Asia and Pacific | 5862 (2168, 11983) | 3462 (1798, 5933) | 33093 (5047, 111697) | 148871 (46042, 348396) | 181964 (68960, 400650) |
| Europe and Central Asia | 549 (197, 1089) | 220 (128, 351) | 2514 (733, 6090) | 8161 (3771, 15647) | 10675 (5529, 19087) |
| Latin America and Caribbean | 608 (269, 1119) | 357 (201, 580) | 1718 (533, 4105) | 8467 (3325, 16451) | 10185 (4485, 18832) |
| Middle East and North Africa | 6.7 (4.5, 9.2) | 4.8 (2.0, 9.2) | 32 (14, 64) | 198 (67, 448) | 231 (94, 479) |
| South Asia | 3597 (1699, 6368) | 2417 (1406, 3873) | 5722 (1121, 15049) | 24383 (8149, 54539) | 30105 (11628, 60923) |
| Sub-Saharan Africa | 7657 (5023, 10857) | 3666 (2266, 5936) | 15097 (7403, 29316) | 110024 (35863, 272481) | 125120 (50295, 291700) |
| Income classification | | | | | |
| Low income | 3555 (2214, 5191) | 1179 (793, 1702) | 2936 (1467, 5030) | 4283 (2475, 7033) | 7219 (4735, 10922) |
| Lower middle income | 7921 (4572, 12520) | 4064 (2551, 6317) | 13797 (6242, 26968) | 44070 (21806, 84397) | 57867 (32965, 99016) |
| Upper middle income | 6804 (2845, 13328) | 4884 (2729, 8006) | 41442 (10334, 121349) | 251751 (102632, 515430) | 293193 (131981, 580979) |
| Time period | | | | | |
| 2000-2009 | 2611 (1613, 3895) | 1315 (846, 1980) | 4514 (2271, 8588) | 27618 (13014, 51929) | 32132 (17718, 57036) |
| 2010-2019 | 9000 (5141, 14426) | 5046 (3125, 7696) | 28326 (11424, 67540) | 152669 (71020, 294430) | 180995 (92207, 334460) |
| 2020-2023 | 6669 (3459, 11681) | 3766 (2244, 5903) | 25334 (8522, 68075) | 119818 (52486, 234829) | 145152 (74070, 273463) |

Values in parentheses represent 95% uncertainty intervals. Cost values represent nominal U.S. dollars. Actual scenario represents the observed scale-up of HIV, TB, and malaria services over the study period. No coverage scenario represents a counterfactual with zero coverage of HIV, TB, and malaria services from year 2000 onward for each country over the study period.

**Table S-4. Sensitivity analysis – Estimates of averted inpatient utilization as a percentage of national hospital bed capacity and averted costs as a percentage of domestic government health expenditure, for actual scenario compared to no coverage scenario, 2000-23.**

| | Number of countries included in analysis | Averted inpatient bed-days (%) | | Averted costs (%) | |
|---|---|---|---|---|---|
| | | Whole period (2000-23) | Final year (2023) | Whole period (2000-23) | Final year (2023) |
| Total | 108 | 6.2 (0.7, 34.3) | 11.0 (1.2, 48.8) | 2.9 (0.1, 8.7) | 3.9 (0.2, 16.1) |
| Disease | | | | | |
| HIV | 108 | 0.7 (0.2, 3.0) | 1.4 (0.4, 5.6) | 0.2 (0.05, 0.8) | 0.3 (0.07, 1.5) |
| TB | 51 | 16.6 (6.1, 30.2) | 27.0 (11.6, 57.6) | 4.5 (2.4, 7.2) | 7.9 (3.5, 14.1) |
| Malaria | 55 | 6.0 (1.5, 11.7) | 5.5 (1.0, 11.9) | 2.9 (0.6, 5.4) | 3.3 (0.7, 6.3) |
| World Bank region | | | | | |
| East Asia and Pacific | 13 | 6.8 (0.8, 13.1) | 11.7 (1.1, 18.7) | 2.4 (0.6, 3.0) | 3.2 (1.0, 4.5) |
| Europe and Central Asia | 13 | 1.9 (0.5, 3.9) | 6.6 (1.2, 10.1) | 2.1 (0.3, 5.4) | 3.5 (0.3, 9.0) |
| Latin America and Caribbean | 18 | 0.8 (0.5, 1.1) | 1.5 (0.9, 2.2) | 0.1 (0.1, 0.2) | 0.2 (0.09, 0.3) |
| Middle East and North Africa | 9 | 0.1 (0.04, 0.14) | 0.2 (0.1, 0.5) | 0.04 (0.02, 0.06) | 0.1 (0.05, 0.2) |
| South Asia | 8 | 12.9 (0.1, 26.5) | 23.4 (0.3, 55.0) | 3.4 (0.04, 6.1) | 5.3 (0.06, 10.0) |
| Sub-Saharan Africa | 47 | 34.7 (11.2, 58.1) | 44.7 (11.8, 96.4) | 9.2 (4.4, 16.6) | 17.2 (5.3, 31.4) |
| Income classification | | | | | |
| Low income | 25 | 35.1 (11.6, 53.3) | 44.7 (11.9, 97.7) | 6.5 (3.5, 16.3) | 9.8 (5.1, 25.2) |
| Lower middle income | 44 | 9.2 (1.0, 34.3) | 14.0 (1.3, 55.7) | 3.5 (0.3, 8.3) | 4.4 (0.6, 16.3) |
| Upper middle income | 39 | 1.1 (0.4, 4.6) | 2.3 (0.6, 10.3) | 0.2 (0.09, 2.4) | 0.3 (0.09, 3.5) |

Point estimates represent median across modeled countries. Values in parentheses represent interquartile range (25$^{th}$ and 75$^{th}$ percentiles) of country-level values.

**Figure S-2. Sensitivity analysis – Partial rank correlation coefficients (PRCCs) for model estimates of averted costs in 2023 for actual scenario compared to constant coverage scenario.**

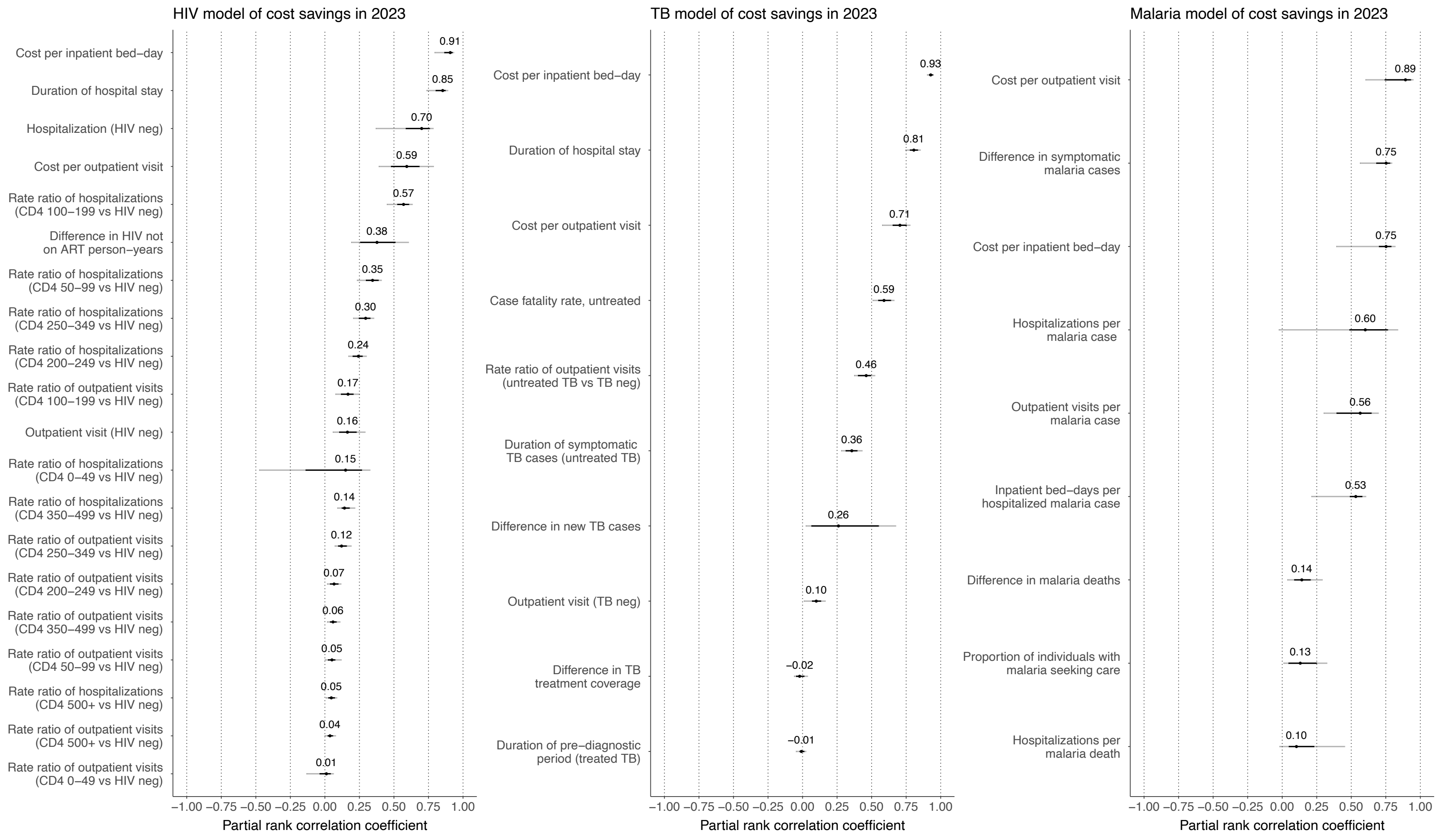

PRCCs estimated for each country individually based on posterior distribution of model parameters. Values (points) indicate median PRCC across countries. Dark lines represent interquartile range of country-level PRCC values. Light lines represent 10th–90th percentiles of country-level PRCC values.

**Table S-5. Sensitivity analysis – Cumulative averted costs due to scale-up HIV, TB, and malaria services under different unit cost assumptions, for actual scenario compared to constant coverage scenario, 2000-23.**

| | Averted costs (US$), millions | | |
|---|---|---|---|
| | Main analysis | Sensitivity analysis: economies of scale | Sensitivity analysis: exceed capacity constraints |
| Total | 134906 (70124, 247531) | 107630 (49532, 207232) | 168153 (78451, 308993) |
| World Bank region | | | |
| East Asia and Pacific | 59375 (22689, 127817) | 47811 (4599, 108990) | 77920 (8097, 175674) |
| Europe and Central Asia | 4327 (2315, 7431) | 3642 (1844, 6239) | 5623 (2985, 9703) |
| Latin America and Caribbean | 5174 (2444, 9764) | 4264 (1650, 8273) | 6141 (2349, 11590) |
| Middle East and North Africa | 197 (80, 421) | 160 (58, 343) | 208 (85, 446) |
| South Asia | 7136 (2781, 14502) | 5499 (2263, 11293) | 8988 (3605, 18174) |
| Sub-Saharan Africa | 58698 (25064, 134770) | 46253 (18592, 106596) | 69272 (29518, 157923) |
| Income classification | | | |
| Low income | 4083 (2673, 6220) | 2487 (1494, 3756) | 4113 (2524, 6075) |
| Lower middle income | 19803 (12533, 31084) | 15147 (9032, 23694) | 24531 (14879, 39667) |
| Upper middle income | 111021 (50911, 222528) | 89995 (33796, 183939) | 139508 (53685, 276619) |
| Time period | | | |
| 2000-2009 | 11795 (6508, 20778) | 8931 (4548, 16048) | 13990 (7469, 24769) |
| 2010-2019 | 72472 (37745, 133518) | 59031 (29632, 110643) | 91910 (47799, 170959) |
| 2020-2023 | 50640 (26167, 91817) | 41361 (20455, 79550) | 64978 (33564, 117474) |

The first sensitivity analysis (economies of scale) allows for unit costs to decline as PHC utilization increases. The second sensitivity analysis (exceed capacity constraints) allows for additional costs at higher levels of utilization, due to the costs of increasing service capacity. Values in parentheses represent 95% uncertainty intervals. Cost values represent nominal U.S. dollars. Actual scenario represents the observed scale-up of TB services over the study period. Constant coverage scenario represents a counterfactual with coverage of TB services held constant at the year 2000 levels for each country over the study period.
